# The Health Interventions Impact Calculator (HIIC): scaling up web-based access to proportional multistate lifetable analyses of avoidable burden, health gain and economic impacts

**DOI:** 10.64898/2026.02.01.26345324

**Authors:** Stephanie Khuu, Tim Wilson, Bibha Dhungel, Samantha Howe, Tony Blakely

## Abstract

Health metrics and modelling capacity have expanded to address present burden and burden attributable to risk factors in the past. There remains a gap in accessible tools that estimate avoidable burden; —that is, the future health and economic impacts of preventive and treatment interventions.

This paper describes and demonstrates the Health Interventions Impact Calculator (HIIC), a free web-based analysis and visualisation tool that allows for rapid estimation of the future health and economic impacts of user-specified intervention scenarios for multiple diseases and risk factors.

HIIC draws on precomputed outputs from the Scalable Health Intervention Evaluation programme (SHINE) proportional multistate lifetable (PMSLT) models. Users define an intervention scenario by specifying intervention timing, target population, intervention cost, and then modifying either disease rates or risk factor exposures. HIIC currently reports outcomes as differences between a business-as-usual (BAU) baseline and the intervention scenario, including health-adjusted life years (HALYs), deaths averted, health system expenditure and income impacts, for Australia. Outputs are presented through interactive dashboard visualisations and downloadable results.

Three example intervention scenarios for Australia are presented: a 10% reduction in ischemic heart disease incidence, a 10% reduction in cervical cancer case fatality rate, and a BMI reduction of 2.5 kg/m² toward the theoretical minimum risk exposure level (TMREL). Across examples, HIIC generates 10-, 20-, and 40-year projections for health gains, mortality displacement over time, and economic impacts. A comparison of interventions based on cost-effectiveness shows how incremental costs and HALYs gained relative to BAU can differ substantially across intervention types, reflecting both intervention design and the level and trajectory of baseline burden.

HIIC is a world-first accessible framework for standardised comparison of intervention scenarios against BAU that will soon be available for all countries. By linking risk factor and disease trajectory changes to health and economic outcomes within a consistent modelling structure, HIIC can inform transparent and reproducible priority setting for decision makers and researchers alike.

**Author Summary:** Decision-makers need to determine which health interventions offer the greatest health gain for the resources invested, but comparing different options has been difficult. While we can measure current disease burden and past impacts, accessible tools for estimating what could be avoided in the future through new interventions have been lacking. We created a free online tool called the Health Interventions Impact Calculator that allows users to explore future scenarios for health interventions. Users enter details about a proposed intervention, such as reducing obesity, improving disease screening, or enhancing treatment, and the tool estimates future outcomes: deaths averted; health improvements, and costs over the next 10, 20, or 40 years. The tool compares each scenario against a business-as-usual future to show what additional benefits the intervention might achieve. We demonstrated this using three examples in Australia: a body-mass index intervention, a heart disease prevention intervention, and improvements to cervical cancer treatment. Currently available for Australia with over 200,000 ready-to-use scenarios and expanding worldwide in 2026, this tool provides researchers, health organizations, and policymakers with a standardised way to rapidly estimate and compare the future impact of different health interventions, supporting evidence-based decisions about where to invest limited resources.

## Introduction

Communities rely on governments and policymakers to cost-effectively invest in policies that improve population health, productivity and support long term prosperity [1–3]. Ageing populations, rising healthcare costs and limited resources make it critical to prioritise interventions based on their expected health, economic and social benefits. However, rapidly and robustly identifying the future benefit of intervention options across prevention, early detection, screening, treatment and rehabilitation, is challenging.

Global health metrics data, data collection, and computational modelling capacity in epidemiology have expanded substantially in the last two decades [4,5]. Large and increasingly granular data sources can be used to communicate model outputs and build data visualisation tools for the public, policymakers, NGOs, and researchers [5]. The challenge now lies in routine and standardised estimation of the health and economic impacts of interventions in future years, and in making these outputs accessible, understandable and actionable for relevant users [6]. Decision support tools in the form of web-based tools and dashboards, such as the Global Burden of Disease (GBD) study Global Health Data Exchange portal for analysing past exposure, have emerged as a solution to bridge this gap between data availability and practical utility [7].

A central source of global health metrics data is the GBD [8], which provides essential burden estimates and counterfactuals, including the health loss or burden in disability adjusted life years (DALYs) across populations, and the attribution of that burden to past risk factor exposure. The original conception of burden of disease studies also aimed to estimate avoidable burden or health gain in the future from public health interventions [8], but this has not been realised at scale.

The Scalable Health Intervention Evaluation program (SHINE) has developed modelling and data infrastructure to support estimating avoidable burden, including disease rate forecasting and intervention models that simulate changes in future health and economic outcomes. In SHINE, interventions can be modelled as direct changes to disease rates (incidence, case fatality, remission and severity), or as indirect changes to risk factor exposures such as tobacco [9] or high body-mass index (BMI) [10] that then translate into downstream changes in disease incidence. The SHINE infrastructure models diseases and priority risk factors simultaneously, improving comparability across interventions and helping to avoid overestimation of health gains that can occur when competing morbidity and mortality are not accounted for.

Many existing digital resources for intervention modelling and comparison are designed for different target audiences and decision contexts and are therefore not directly comparable across interventions and populations. Some tools are disease-specific, others draw on methodologically heterogeneous inputs, or not widely accessible, and many are not designed for head-to-head comparisons across a broad set of disease and risk factor interventions, across populations [11,12]. For example, FairChoices [13] is oriented toward priority setting in low-and middle-income country contexts based on a library of existing interventions, whereas WHO-CHOICE [14] provides generalised cost-effectiveness evidence intended to support health benefit package design, and the Disease Control Priorities project [15] synthesises evidence across multiple studies and settings. While each of these resources is valuable for its intended purpose, differences in scope, target populations, and underlying assumptions mean they do not provide a single, consistent framework for comparing intervention scenarios against a common business-as-usual baseline within one modelling structure.

To make modelling output widely available for decision support, SHINE has created the Health Interventions Impact Calculator (HIIC)—a world first data analysis and visualisation web tool that estimates outcomes from user-specified intervention scenarios across key risk factors and more than 150 diseases. HIIC reports outputs from SHINE’s proportional multistate lifetable (PMSLT) models, showing differences between a business-as-usual (BAU) baseline and a user-selected intervention scenario.

The HIIC interface currently provides users free access to more than 200,000 precomputed intervention scenarios for Australia and will soon be deployed for all countries. Users can input theorised intervention parameters within the calculator panel (Figure 1) to generate estimates of health and economic outcomes. In disease-focused interventions, users can modify incidence, severity, case fatality, and remission rates. For risk factor interventions, users can specify absolute or percentage changes in exposure toward the theoretical minimum risk exposure level (TMREL). The platform reports projected health gains, deaths averted, and economic outcomes, including cost savings and cost-effectiveness. By combining large-scale PMSLT outputs with user-defined adjustments to risk factor and disease trajectories, HIIC supports rapid, early-stage scoping of potential health and economic impacts relative to baseline scenarios. HIIC is a valuable digital tool that may help inform complex decision making in research and government priority setting.

**Figure 1.**
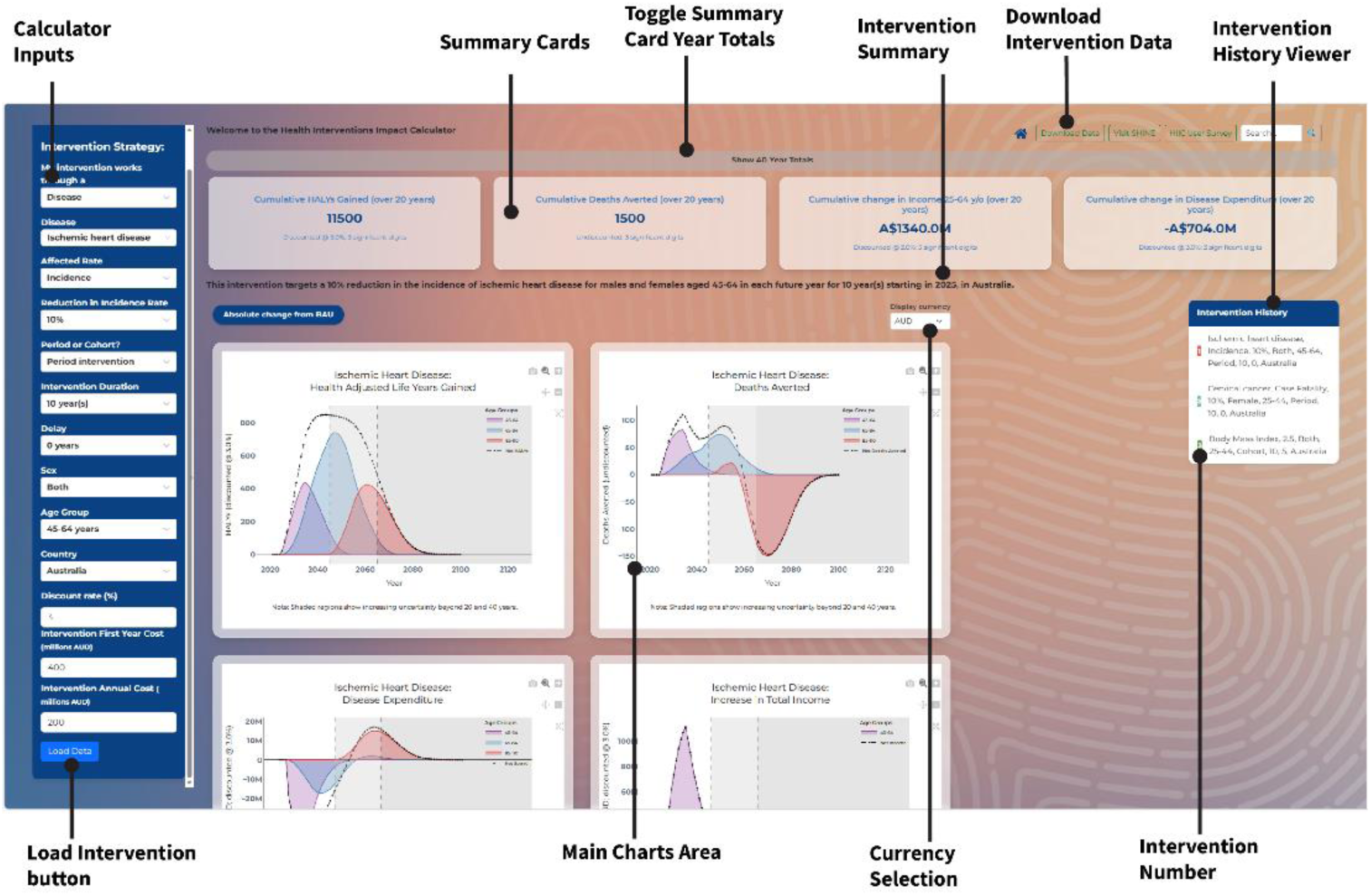
Annotated screenshot of the HIIC dashboard showing the main interface components: calculator inputs (left panel), summary cards and toggleable totals (top row), intervention summary and download controls (top right), intervention history viewer and intervention identifier (right), main chart area (centre), and the “Load intervention” button (bottom left).

This paper describes the framework underpinning HIIC, defines the input parameters for the webtool, and illustrates its use through three example applications in Australia: reduction in ischemic heart disease incidence, reduction in cervical cancer incidence, and body mass index (BMI) reduction toward the theoretical minimum risk exposure level (TMREL).

## Methods

Key elements and assumptions required for users to understand example intervention scenario outputs via the HIIC tool are summarised below; detailed PMSLT methods are described by Blakely et al 2020 [16]. Video tutorial, tutorial IHD example and documentation for HIIC can be found at https://shine-hiic.com/docs.

### Modelling approach

#### Framework

Governments and statistical agencies commonly use lifetables to estimate period life expectancy: the expected life years a population would live if experiencing age-specific mortality rates in a given calendar year. Less commonly, cohort lifetables are used to estimate a birth cohort’s life expectancy given the rates they experienced (or are forecast to experience) as they age. PMSLT modelling applies a cohort approach to estimate both business-as-usual (BAU) and intervention expected life expectancy, life years and other outcomes, with health gain defined as the difference between BAU and intervention outputs.

As a hybrid lifetable and Markov state-transition model, typically implemented with annual cycles, PMSLT simulation can attach state weights such as morbidity and costs and accumulates these over time. This enables estimation of combined mortality and morbidity metrics, e.g. health-adjusted life years (HALYs) gained, and economic impacts such as changes in health system expenditure on disease.

In PMSLT simulation (Figure 2), disease lifetables are modified through intervention scenarios that alter disease incidence, severity, case fatality, or remission rates, which in turn affect population-level mortality and morbidity rates. This can be used to compare multiple output metrics, namely HALYs, life expectancy, and health system expenditure, between a business-as-usual baseline and intervention scenario. The comparison occurs for a user-determined time horizon (e.g. the next 20 years).

**Figure 2.**
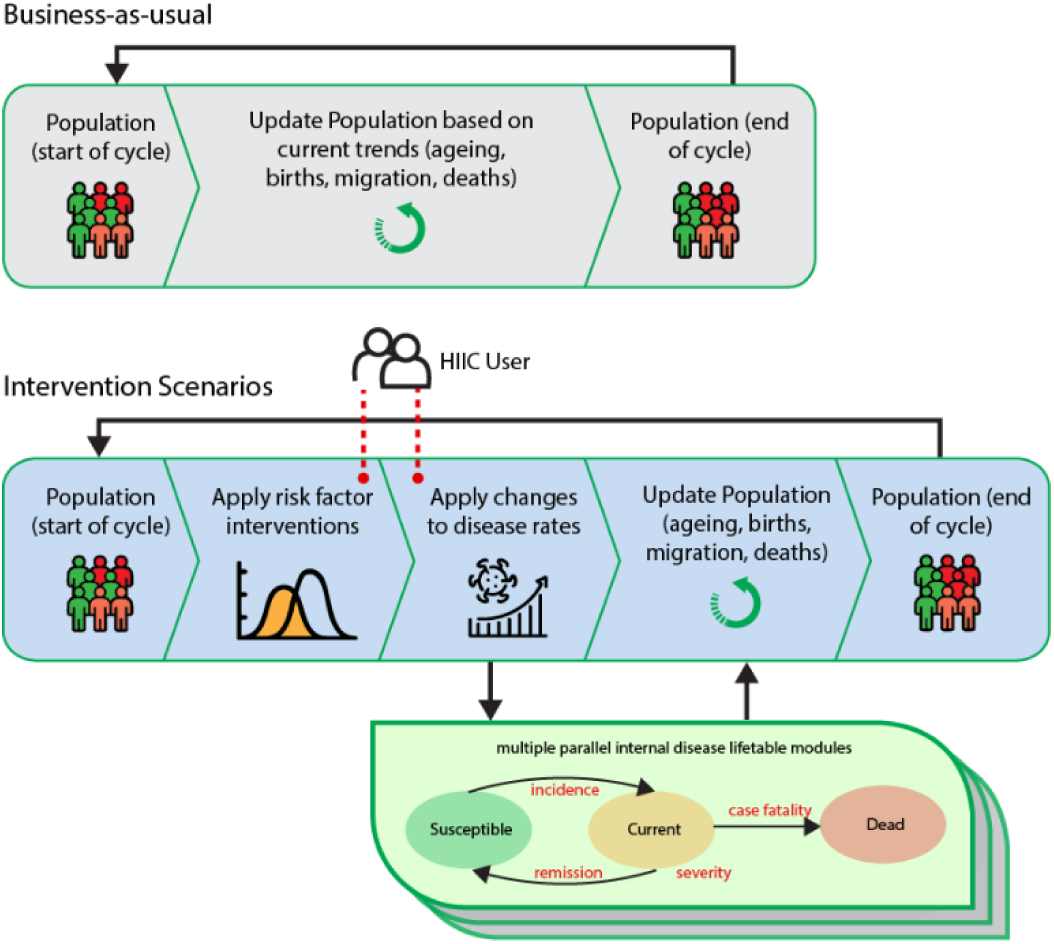
Conceptual overview of PMSLT modelling workflow. User-specified interventions on HIIC alter a single risk factor that flows on to effect incident rates in all causally associated diseases. Disease interventions can alter disease rates (incidence, case fatality, remission or severity). Both impact population level mortality and morbidity.

#### Data sources and input specifications

HIIC is currently deployed with data for Australia but will soon incorporate estimates for all countries included in the GBD study. For each country, inputs are parameterised by sex and age, using forecasts for demography, disease epidemiology rates (future incidence, case fatality and remission), all-cause mortality and morbidity, and risk factor exposure distributions. Disease rates and all-cause mortality forecasts use GBD rates as inputs [17]. These rates are then calibrated to ensure cohort-coherent historical data and forecasts [18]. For disease and all-cause morbidity, we use the GBD years of life lived with disability divided by population counts and assume these rates remain constant into the future. Risk factor forecasts are generated by applying an estimation method to derive risk factor distributions from GBD summary exposure value forecasts [19].

Health system expenditure estimates, including costs in the first year of diagnosis, last year of life if dying from that disease, and prevalent disease years, is assigned to states in the model. [20]. When HIIC expands to all countries, this component will be available for OECD countries.

For Australia and New Zealand, income loss by disease phase [21] is likewise assigned to states in the model.

#### Population and horizon

Results follow the PMSLT base year (2025) and time horizon for the chosen country. HIIC supports two implementation approaches. In a cohort approach, the parameter change is applied to the baseline cohort within a specified age band and follows this cohort as they age. In a period approach, the change is applied to the eligible population in each future year, with individuals ageing into and out of eligibility over time.

#### Time lags for interventions acting through risk factor change

Time lags refer to the delay between a change in risk factor exposure and the resulting change in disease risk and incidence. For most risk factors, disease risk does not respond immediately to exposure changes, and the length of this delay varies by disease type. As a result, estimates of avoidable burden must account for the fact that current and future disease risk depends partly on past exposure.

In SHINE, time lags for tobacco related diseases use decay functions since quitting from Hoogenveen et al [22]. For other risk factors time lags are incorporated with a generic method that takes the weighted average of potential impact fractions (PIF) from previous years, i.e. current year disease risk is a function of lagged risk factor exposure. Three generic lags are used: long duration of two to 22 years of past or look-back window exposure (mainly cancers); short duration of last five years of exposure (mainly cardiovascular disease); and nil or current exposure (i.e. acute diseases like lower respiratory tract infection). As shown in panel B of Figure 3, the PIF used is a weighted average of PIFs in each year within the look-back window.

**Figure 3.**
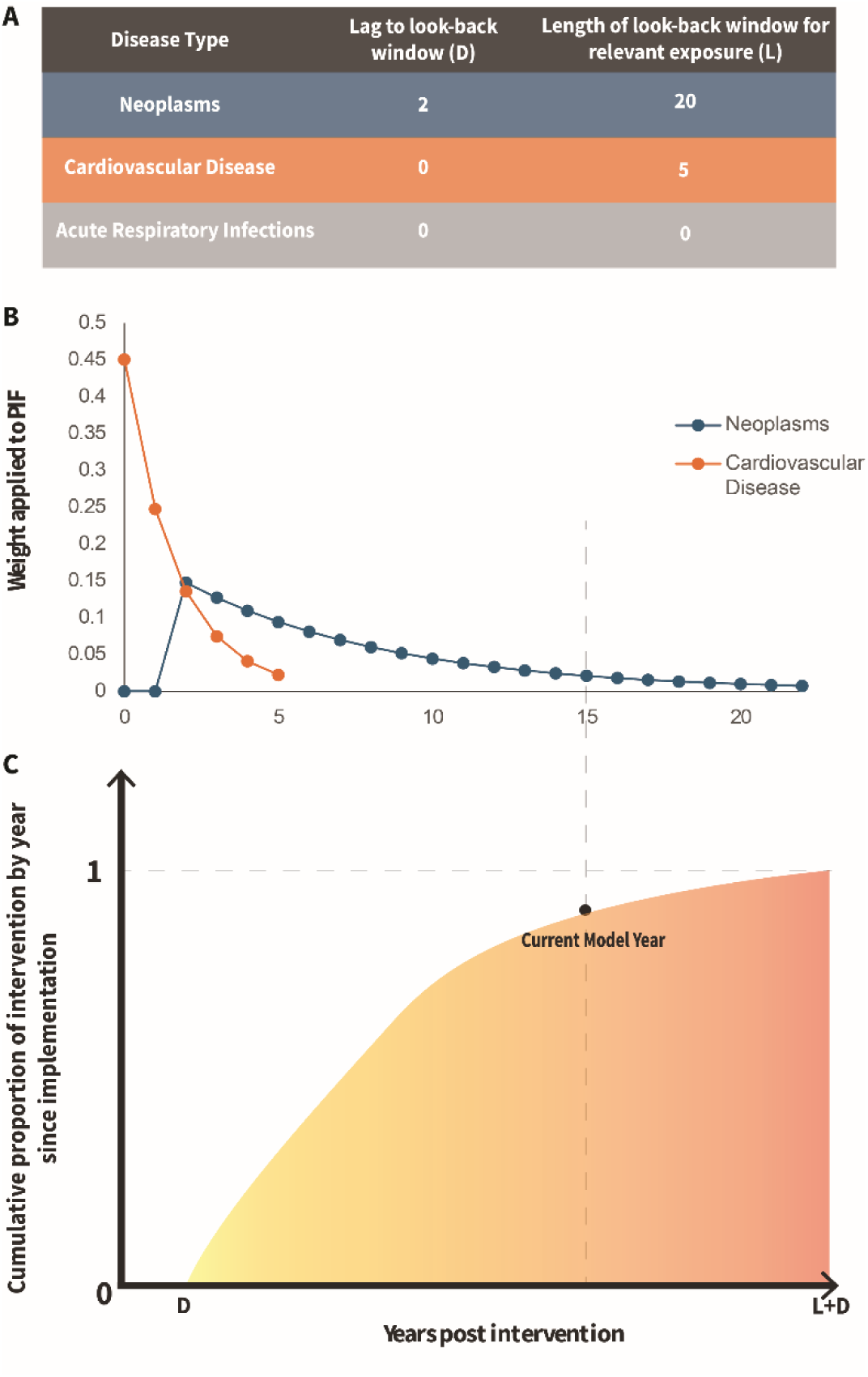
Risk factor to disease time lags for different disease types. B) The weight applied to the PIF with years post intervention, and C) the cumulative proportion of intervention effect size applied to the disease incidence since implementation, including the delay (D) and lag (L).

#### The scenarios

##### Business as usual (BAU)

The BAU scenario represents a counterfactual in which no new interventions are introduced and existing behaviours, exposures, and policies remain unchanged. Disease rates follow their recent trajectory for a defined projection period of 20 years; rates are then held constant to avoid implausible long-term extrapolation.

##### Intervention

a modelled counterfactual or “what-if” version of the future assuming a specific policy or program is put in place either through changes in a risk factor distribution, or in a disease rate. The model then updates population risk factor exposures and disease rates based on that change. HIIC compares this intervention future to the business-as-usual future. The difference represents the estimated intervention effect on health and economic outcomes.

#### Rates and exposure (HIIC modes)

In disease mode, users modify one disease parameter at a time (incidence, remission, mortality/case fatality, or severity). In risk factor mode, users modify a risk factor exposure distribution toward the TMREL, either by 1) an absolute shift in the native exposure unit or 2) a percentage movement. Changes in exposure are mapped to changes in disease incidence using the PIF approach [8].

#### Theoretical Minimum Risk Exposure Level (TMREL)

The risk factor exposure level at which population risk is lowest. A percentage reduction for a risk factor moves the exposure distribution toward this optimum.

#### Discounting

When a discount rate is specified, yearly outcomes for HALY and cost outputs are discounted prior to calculating total values displayed in summary outputs and downloads (deaths averted are not discounted). Present value, 𝑃, is calculated as 𝑃 = 𝑉_𝑛_ / (1 + 𝑟)^𝑛^, where 𝑉_𝑛_ is the outcome value 𝑛 years from the base year and 𝑟 is the discount rate.

#### Health and economic outcomes

HIIC uses precomputed PMSLT outputs derived from thousands of intervention scenarios applied to a given country. Health-adjusted life years (HALYs) are accumulated life years adjusted for morbidity loss. Deaths averted represent the difference in the timing of deaths between BAU and intervention scenarios. Over a complete lifetime horizon, total deaths in BAU and intervention are the same (as everyone has a probability of 1 of dying); but an intervention delays deaths to older ages, with deaths averted being the difference in deaths in that year (or cumulative to that year) between intervention and BAU.

Annual differences in health system expenditure are estimated by estimating disease-specific spending under the intervention scenario and summing across sex-age strata. Results are reported as net differences relative to BAU. For Australia, income impacts are estimated using phase-specific income loss weights as described above. Currency conversions are applied using CPI (index 2010) for desired currency in 2024 and 2024 PPP from World Bank Data Bank [23].

HIIC currently provides access to more than 200,000 precomputed intervention scenarios for Australia based on PMSLT outputs and will in early 2026 provide similar numbers of estimates for all other countries. Users select an intervention and adjust disease or risk factor inputs within supported modes. The platform then displays projected differences in health and economic outcomes relative to BAU, including HALYs gained, deaths averted, changes in health system costs, and cost-effectiveness metrics where available. Outputs are presented through interactive visualisations and summary cards, with downloadable results for further analysis. Here, cumulative gain is summarised over 20 years, with possible options of 10, 20 and 40 years. All output monetary values for the following examples are reported in 2024 USD.

#### Example Intervention Scenarios

A policymaker is developing a business case for expanding structured chronic disease checks in primary care (e.g., through enhanced Medicare item use and practice incentives) and requires estimates of health gain. The intervention is assumed to operate for 10 years from 2025 and is hypothesised to reduce the incidence of ischemic heart disease (IHD) by 10% amongst males and females aged of 45-64 yrs during the implementation period. Programme costs are set at AUD 400 million (USD 293million) in the first year (start-up, training and rollout) and AUD 200 million USD 146 million) per year thereafter (ongoing delivery). Outcomes are discounted at 3%.
A researcher is designing an evaluation of an intensification of cervical cancer follow-up (e.g., targeted improvements in participation, timeliness of follow-up, and treatment pathways) and requires estimates of the expected impact. The intervention is modelled as a period programme program implemented annually for 10 years from 2025 for females aged 25-44-years, with the effect represented as a 10% decrease in the cervical cancer case fatality rate during the intervention period. Costs are set as AUD 100 million (USD 73.2 million) per year, with first year costs of AUD 157 million (USD 115 million). The same discount rate of 3% is applied.
An NGO, commissioned through a government partnership and coordinating healthcare delivery across primary health networks and community providers, is developing a large-scale, targeted weight management programme and requires estimates to inform funding applications and evaluation planning. The intervention commences in 2030 and is implemented for 10 years for a cohort of males aged 25-44 years, with the estimated effect of reduction in BMI by 2.5kg/m^2^ towards the TMREL of 22.5kg/m^2^, for those who are overweight or obese. Programme costs are set at AUD 500 million (USD 366 million) in the first year with 300 million (USD 220 million) annual recurring costs. Outcomes are discounted at 7%.

## Results

HIIC dashboard and data download functions were used to generate visualisations for the three example interventions. All intervention scenarios posed for Australia provide HALYs, deaths averted, disease expenditure, and the increase in total income as outputs. Values are rounded to 3 meaningful digits.

Over 20 years, the IHD intervention generated a cumulative gain of 11,500 HALYs, 1,500 deaths averted, an estimated USD 970 million (AUD 1340 million) in total income for 25–64-year-olds, and a cumulative change in disease expenditure of USD -509 million (net savings; AUD 704 million) (Figure 4). The cervical cancer intervention, which applies to females at younger ages and targets a less common disease, produced much smaller health and economic impacts: cumulative 20-year changes of 394 HALYs gained, 44 deaths averted, an estimated USD 17.5 million (AUD 24.1 million) increase in total income, and a cumulative increase in expenditure of USD 2.6 million (AUD 3.6 million). This increase in expenditure is consistent with interventions that reduce case fatality rates alone, which can increase the prevalent pool of disease and therefore increase health system expenditure. The BMI reduction intervention, although commencing five years later, produced substantial impacts because BMI is a common risk factor, is forecast to increase in the population under BAU, and affects many diseases. It generated 60,800 cumulative HALYs gained over 20 years, 3,200 deaths averted, an estimated USD 3.06 billion (AUD 4.10 billion) increase in total income, and USD 1.14 billion (AUD 1.53 billion) in cumulative savings in health expenditure.

**Figure 4.**
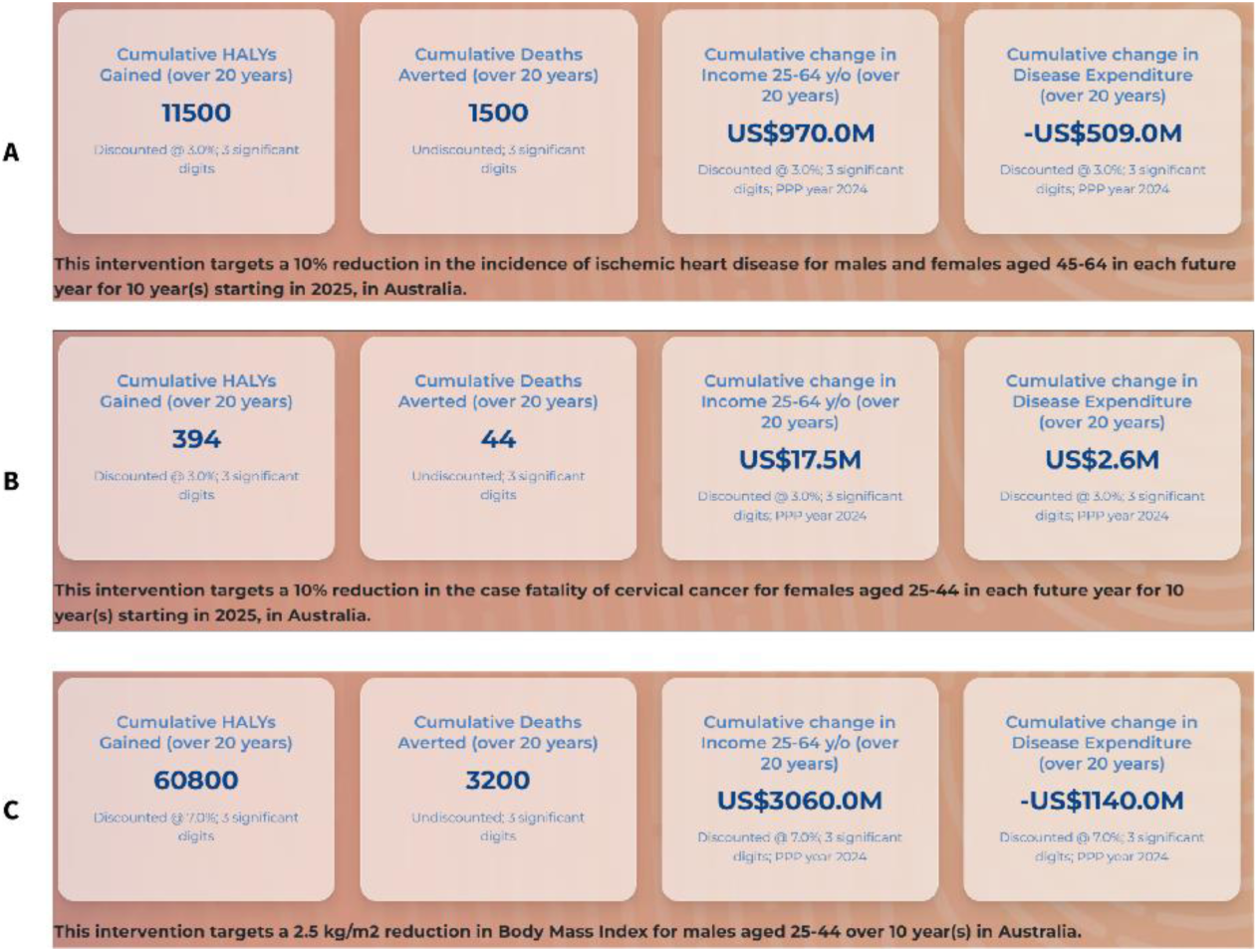
Screenshots of cumulative HALY gain, deaths averted, income gain and health savings over 20 years for example intervention scenarios A (ischemic heart disease), B (cervical cancer), and C (body-mass index).

Figure 5 shows HIIC dashboard outputs for the IHD intervention. Whilst the intervention targets people aged 45–64 years, and HALY gains were substantial in this age range, the largest annual HALY gain occurred at older ages (65–84-years), peaking in 2047 (741 HALYs). This propagated health gain reflects deaths being averted at younger ages (before 45-years), with additional life years accruing as the cohort survives into older age, which more than offsets the return to BAU incidence rates post age 45. Deaths averted (Figure 5B.) were positive in early years but became negative in later years (i.e. more deaths occurring under the intervention scenario) as the population aged into the 85–100-year-olds group, reflecting deaths being delayed to older ages under the intervention. A similar pattern was observed for disease expenditure: early cost savings concentred in ages 45–64 years and increasing future expenditure for older age groups (85–110 years) (Figure 5C).

**Figure 5.**
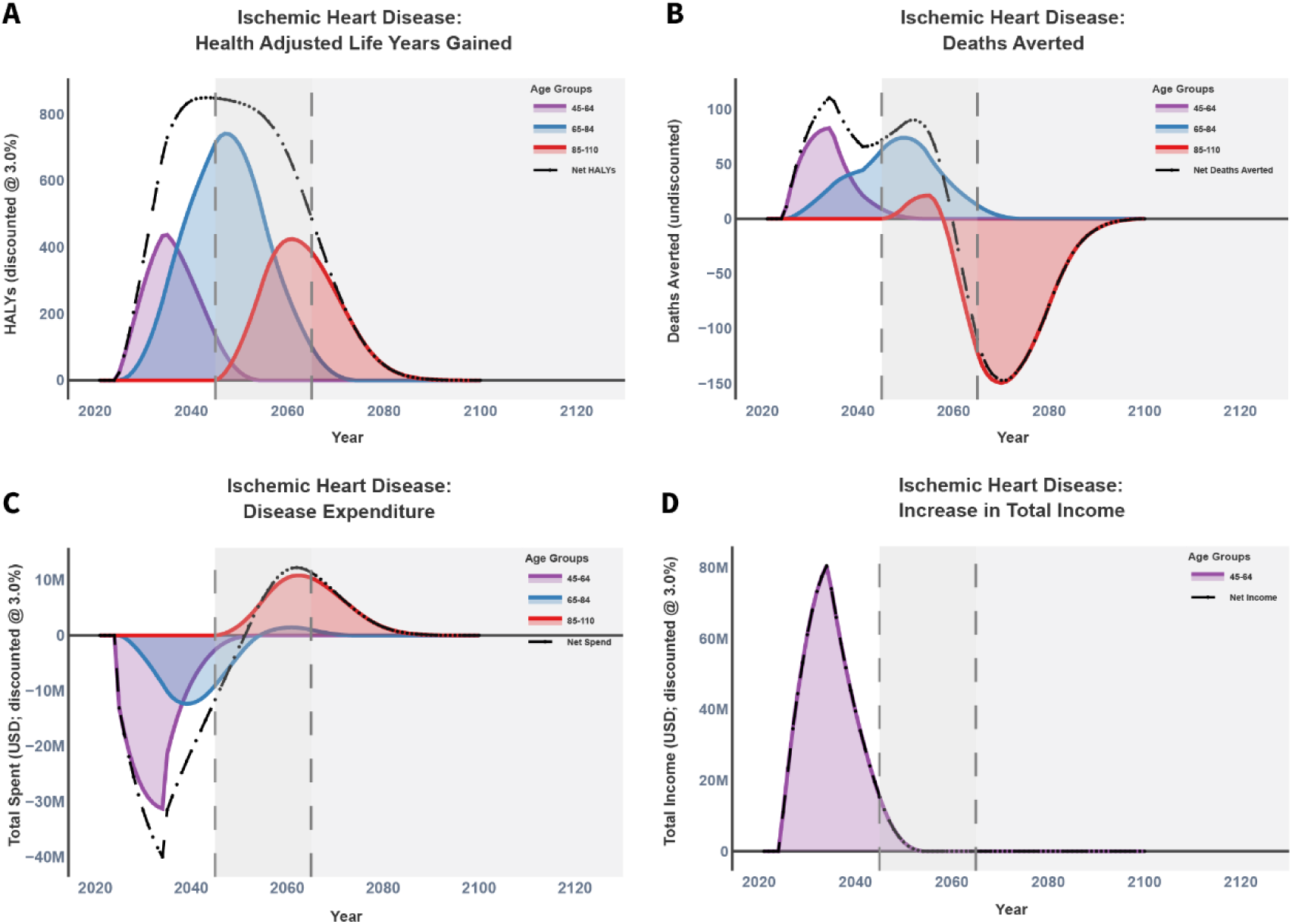
HIIC Dashboard results for a 10-year period intervention with 10% reduction in incidence of ischemic heart disease for males and females aged 45-64 in Australia. A) HALYs: Health adjusted life years. B) Deaths averted C) Disease Expenditure, and D) Increase in total income for 45–64-year-olds. Grey and dark grey areas show increasing uncertainty.

Figure 6 shows the cervical cancer case fatality rate intervention. Peak annual HALY gain occurred in 2035 with 25-44-year-olds and 45-44-year-olds generating 27 HALYs for the cervical cancer intervention (Figure 6A). A peak of five deaths per year were averted in the 25-44-year-old category. There were cost savings seen in the first year of the intervention due to avoiding end of life expenses for some women, followed by increased expenditure due to both an increasing prevalent pool of women living with cervical cancer, and an increasing cured pool living without cervical cancer but incurring other health expenditure. The peak increase in net income gain is also observed in the last year of the intervention (2034; USD 1.21 million).

**Figure 6.**
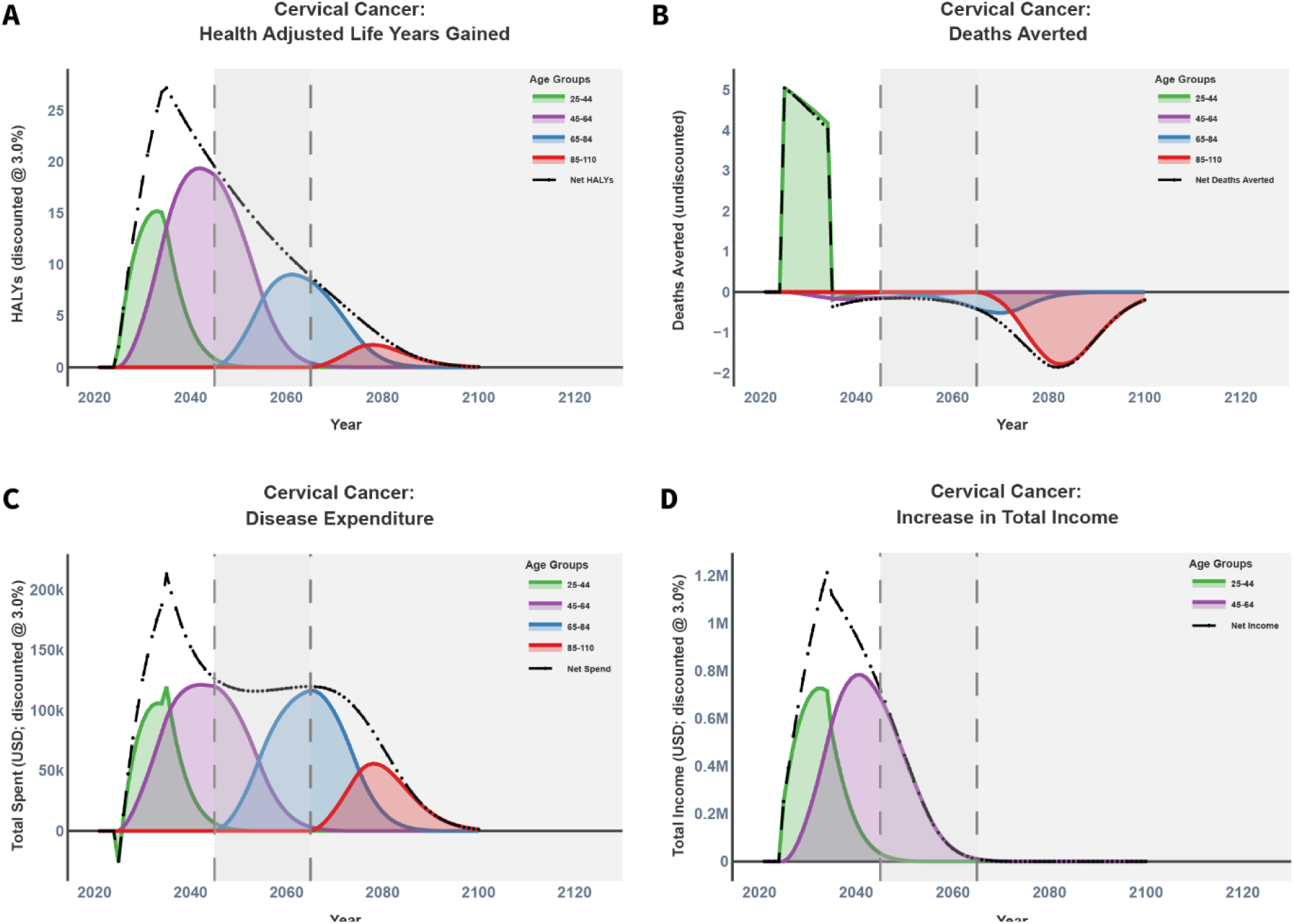
Dashboard results for a 10-year period intervention with 10% reduction in case fatality of cervical cancer for females 25-44 in Australia. HALYs: Health adjusted life years. Grey and dark grey areas show increasing uncertainty.

The BMI intervention produced peak annual HALY gains occurred in 2040 (6,152 HALYs gained in that year; Figure 7), primarily among ages 45–64 years. This pattern reflects both the propagated health gains from lives saved before the age of 45 and, more importantly, the time lags in the modelling, whereby chronic disease incidence is driven by exposure 5 to 22 years (median) in the past (Figure 3). Note that the BMI reduction only occurs for ages 25 to 44 years; upon turning 45 years, the cohort reverts to the BAU BMI distribution. Deaths averted remained positive until approximately 2070, after which they became negative as the cohort aged into the 85–110-year group, again reflecting delayed mortality rather than permanent prevention. Health system expenditure remained net saving until 2062, peaking in 2040 (net savings USD 113 million). The annual increase in total income peaked at USD 382 million in 2041.

**Figure 7.**
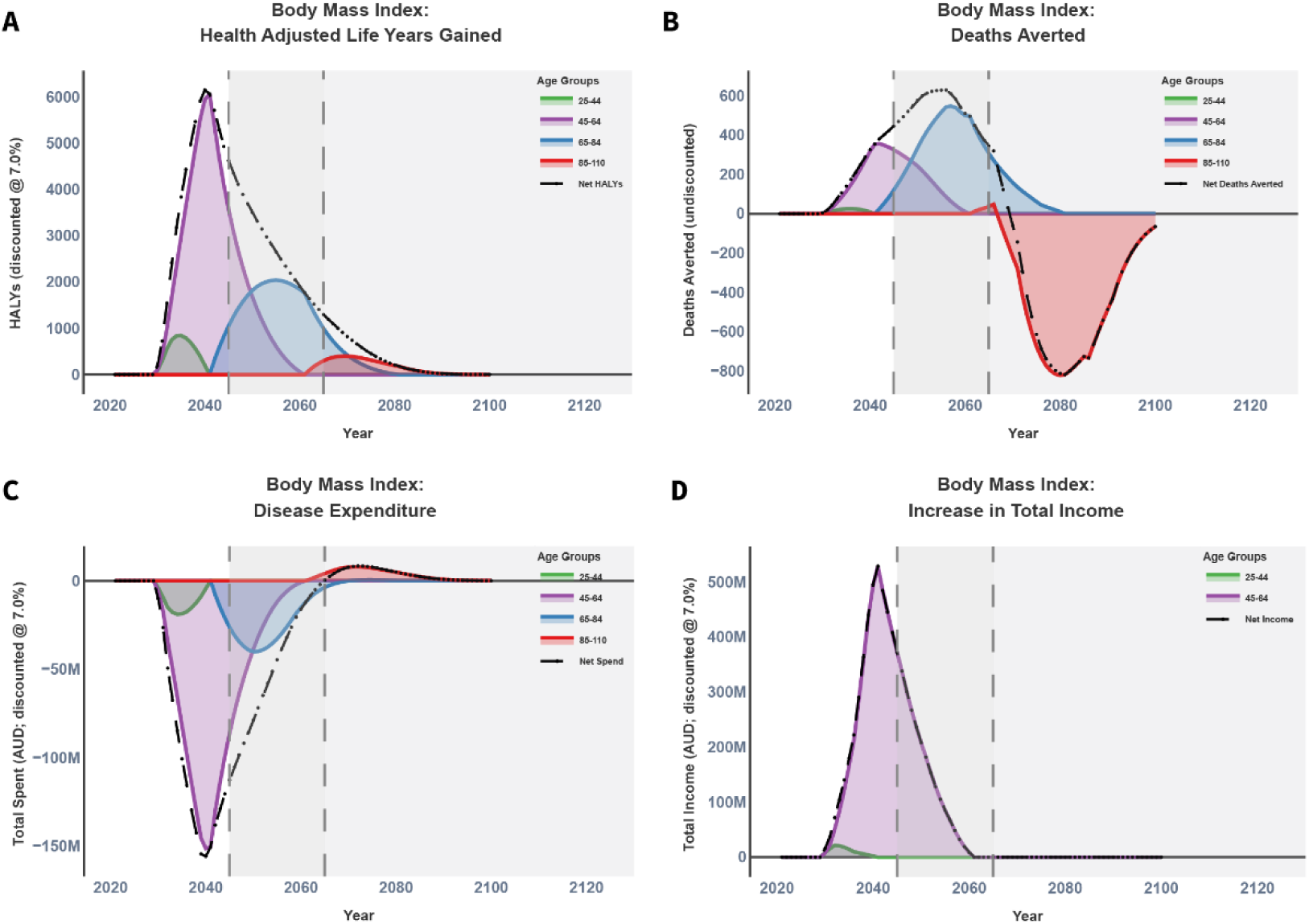
BMI reduction by 2.5kg/m^2^ towards TMREL for a cohort of males between the age of 25-44 years old for 10 years.

Figure 8 below summarises incremental cost-effectiveness over a 20-year horizon by plotting incremental costs (the net of intervention costs entered by the user, and downstream changes in health system expenditure estimated by the PMSLT) against incremental health gains (HALYs) for each intervention, relative to BAU, with the dashed line indicating a willingness-to-pay threshold of USD 50,000 per HALY gained. The BMI intervention lies well below the threshold line, with an ICER of USD 11,394 per HALY, indicating it is highly cost-effective under this threshold given its large health gains for relatively low net cost and even for a discount rate of 7% (its ICER at a discount rate of 3% was USD 2,510 per HALY). In contrast, the ischaemic heart disease intervention (ICER USD 83,075 per HALY) and the cervical cancer intervention (ICER USD 1,780,101 per HALY) fall above the threshold line, indicating they are not cost-effective at USD 50,000 per HALY in these example specifications, with the cervical cancer scenario in particular reflecting comparatively small HALY gains for a positive incremental cost. However, a policy maker would often want to see a longer time horizon for cost effectiveness: for a 40-year time horizon (and same discount rate of 3%) the ICERs were US$38,630 per HALY and US$ 1,042,583 per HALY for the IHD and cervical cancer interventions, respectively.

**Figure 8.**
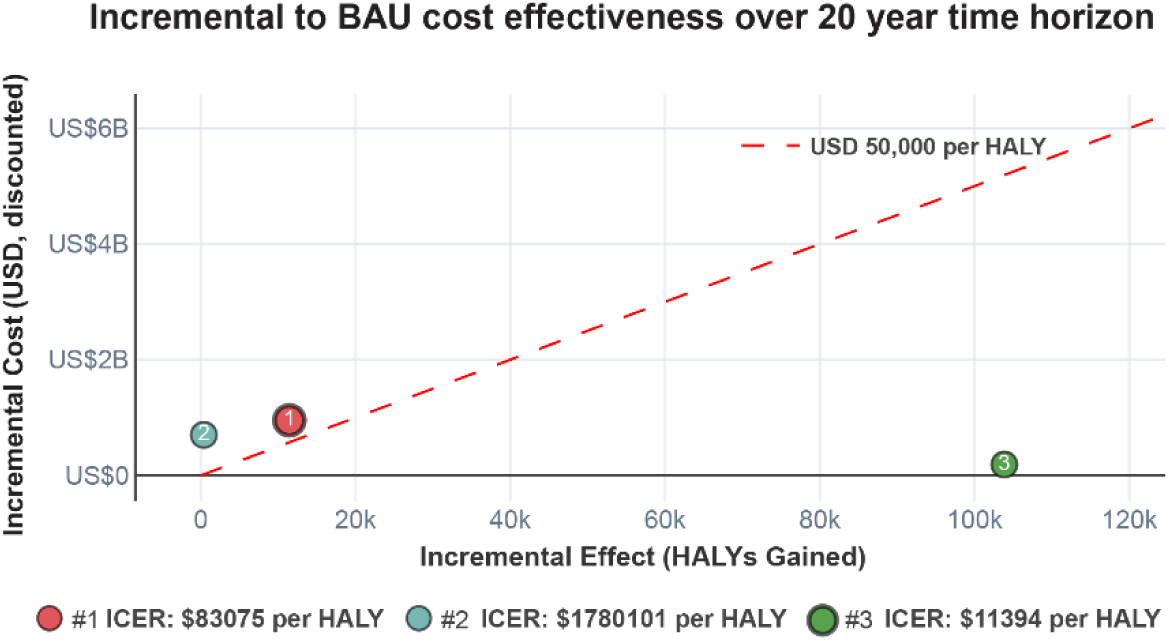
Incremental to BAU cost-effectiveness plot for (1) ischemic heart disease (discount rate 3%), (2) cervical cancer (discount rate 3%) and (3) BMI intervention (discount rate 7%), respectively. The willingness to pay amount (red dashed line) was set to USD 50,000.

## Discussion

HIIC provides a world-first data visualisation tool of intervention scenario health and economic impacts, in any country, to help prioritise and optimise interventions based on projected health benefits and economic outcomes. By enabling rapid and standardised comparison of intervention scenarios across diseases and risk factors, HIIC can support the development of health intervention research and policy briefs with more timely, standardised evidence. In this way, HIIC may assist researchers, NGOs and policymakers to identify high-value interventions under constrained budgets, with potential reductions in health system expenditure and improvements in workforce productivity via estimated income gain.

Existing decision-support resources serve different purposes and targets, and differences in scope, accessibility, and assumptions limit direct head-to-head comparison across a wide range of interventions. Tools such as FairChoices, WHO-CHOICE and Disease Control Priorities provide valuable synthesis and generalised cost-effectiveness evidence, while other platforms offer disease- or setting-specific modelling. HIIC’s contribution is the combination of multi-disease outputs, consistent structure for scenario comparison, and a free interface that links risk factor and disease trajectory changes to health and economic outcomes.

Interpreting the cervical cancer results also highlights the importance of comparing interventions to a BAU baseline rather than to a static “no prevention” counterfactual. In Australia, BAU already reflects substantial ongoing reductions in cervical cancer driven by long-standing HPV vaccination and the National Cervical Screening Program, alongside an explicit national goal of eliminating cervical cancer as a public health problem by 2035. As a result, projected future burden in Australia under BAU is already low (e.g., incidence around 6.6 per 100,000 women in 2020, approaching the WHO elimination threshold of <4 per 100,000) [24], leaving limited avoidable burden for additional interventions to avert in an average-population scenario. However, a HIIC analysis for cervical cancer in other countries could be very different.

Key limitations of HIIC include the scope of conditions and interventions currently represented, and reliance on available epidemiological, cost and income data. Outputs should be interpreted as model-based estimates for early-stage scenario scoping rather than definitive evaluations (e.g. a policy maker may require simultaneous changes to colorectal cancer incidence, case fatality and severity for a program like screening —a functionality not yet provided by HIIC). HIIC results will vary with assumptions about intervention effect size, timing, and implementation approach.

A future direction to improve HIIC includes using machine learning prediction, trained on PMSLT outputs, to allow users to request many more intervention options (i.e. more than the current pre-run options) and to access packages of interventions (e.g. including all possible dyads of risk factor interventions, such as a BMI and blood pressure reduction program together, would be too cumbersome through pre-run models).

From an implementation science perspective, adoption in practice will depend on usability, digital literacy, and internet access, alongside stakeholder engagement and alignment with decision-making processes. To reduce the risk of misinterpretation, HIIC includes embedded guidance at the point of use, including definitions of outputs, prominent labelling of results and description of key modelling settings within user documentation. HIIC has also been developed with user-centred design approach to support usability and uptake. An ongoing co-design survey (<u>hiic-survey.shinyapps.io/hiicsurvey/</u>) has been implemented to capture feedback from intended users (e.g., researchers and policymakers) on usability, interpretability of outputs, and priority features for future development. In parallel Google Analytics (GA4) is used to monitor real-world engagement with the platform, showing 963 active users across 7 countries in the first four months after the webtool was launched. Together, these data sources inform iterative refinement of the interface, documentation and training materials. A follow-up study that formally evaluates HIIC will be used to support development and design decision-making over time.

HIIC provides an accessible decision-making framework to support prioritisation and optimisation of interventions based on projected health benefit and cost-effectiveness. By enabling faster identification of high-value options, HIIC may contribute to stronger evidence for policy and investment decisions, potential reductions in health expenditure, and improvements in population health and productivity.

## Data Availability

All relevant data are available from the HIIC website, www.shine-hiic.com

## Acknowledgements

We acknowledge the provision of disease rate and population data provided by IHME (Source: Institute for Health Metrics Evaluation. Used with permission. All rights reserved.).

